# Loss of Y in regulatory T lymphocytes in the tumor micro-environment of primary colorectal cancers and liver metastases

**DOI:** 10.1101/2023.06.17.23289722

**Authors:** Magdalena Wójcik, Ulana Juhas, Elyas Mohammadi, Jonas Mattisson, Kinga Drężek-Chyła, Edyta Rychlicka-Buniowska, Bożena Bruhn-Olszewska, Hanna Davies, Katarzyna Chojnowska, Paweł Olszewski, Michał Bieńkowski, Michał Jankowski, Olga Rostkowska, Andrzej Hellmann, Rafał Pęksa, Jacek Kowalski, Marek Zdrenka, Jarek Kobiela, Wojciech Zegarski, Wojciech Biernat, Łukasz Szylberg, Piotr Remiszewski, Jakub Mieczkowski, Natalia Filipowicz, Jan P. Dumanski

**Author notes:** Magdalena Wójcik, Ulana Juhas and Elyas Mohammadi contributed equally to this paper and share the first authorship. Natalia Filipowicz and Jan P. Dumanski share the senior authorship.

## Abstract

Male sex is a risk factor for colorectal cancer (CRC) with higher illness burden and earlier onset. Thus, we hypothesized that loss of chromosome Y (LOY) in the tumor micro-environment (TME) might be involved in oncogenesis. Previous studies show that LOY in circulating leukocytes of aging men was associated with shorter survival and non-hematological cancer, as well as higher LOY in CD4+ T-lymphocytes in men with prostate cancer vs. controls. However, nothing is known about LOY in leukocytes infiltrating TME and we address this aspect here. We studied frequency and functional effects of LOY in blood, TME and non-tumorous tissue. Regulatory T-lymphocytes (Tregs) in TME had the highest frequency of LOY-cells (22%) in comparison to CD4+ T-lymphocytes and cytotoxic CD8+ T-lymphocytes. Using scRNA-seq LOY was also linked to higher expression of *PDCD1, TIGIT* and *IKZF2* in Tregs. *PDCD1* and *TIGIT* encode immune checkpoint receptors involved in the regulation of Tregs function. Our study sets the direction for further functional research regarding a probable role of LOY in intensifying features related to the suppressive phenotype of Tregs in TME and consequently a possible influence on immunotherapy response in CRC patients.

## Introduction

Colorectal cancer (CRC) is one of the leading contributors to cancer-related morbidity and mortality worldwide [1]. The liver is a main site of metastases from primary tumors of CRCs, likely due to direct blood drainage through portal system. Consequently, about 50% of CRC patients develop synchronous or metachronous liver metastases (LM_CRC) [2], which frequently leads to the death of the patient [3]. Observations of CRC cohorts conducted for over the past 25 years indicate that CRC shows pronounced sexual dimorphism [4]. Male sex is a risk factor for CRC, with higher illness burden, and earlier onset and metastatic disease, of which ∼30% are synchronous with CRC diagnosis [4]. Mutations in numerous genes have been associated with CRC risk, development of recurrence and metastatic disease [5]. However, there is limited evidence that could explain the higher risk and worse prognosis of men with CRC compared to women, and this aspect attracted our attention because of possible role of the mosaic loss of chromosome Y (LOY) in the disease process [6].

LOY is the most frequent human post-zygotic mutation and is most prevalent in the hematopoietic system of aging men. It is detectable in whole blood DNA from >40% of men above the age of 70 years [7], reaching 57% in the analysis of 93-year-old men [8]. Recent single-cell transcriptomic analyses of leukocytes from 29 aging men (median age 80 years) identified cells with LOY in all studied subjects [9]. LOY has also been observed in other tissues, but with lower frequencies [8, 10]. The most recent example of LOY analysis in another organ than the hematopoietic system was performed in human brains [11]. Serial analysis of men showed that LOY is a dynamic process [12, 13]. LOY presumably causes a clonal expansion of affected cells and may coexist with so called CHIP mutations [14]. Moreover, LOY affects different lineages of hematopoietic cells with varying frequencies and it plays a role in transcriptional dysregulation of autosomal genes in a pleiotropic, cell type specific manner [9]. Furthermore, dysregulation of immune genes was pronounced in cells with LOY, particularly related to immune checkpoint (IC) genes [9, 15, 16]. Major risk factors for LOY include age, smoking and germline predisposition [7, 8, 17, 18]. LOY has been associated with increased risk for all-cause mortality and many common age-related diseases, predominantly chronic, but also acute, inside and outside of the hematopoietic system. Notably, LOY likely has an impact on susceptibility to cancer [6, 7], Alzheimer’s disease (AD) [17], cardiovascular diseases [19], and severity of COVID-19 [13]. Previous studies suggested that LOY disrupts proper functions of the immune system, especially the immune-surveillance mechanisms in Alzheimer’s disease and cancer [9, 16]. Moreover, patients with prostate cancer showed higher levels of LOY in granulocytes and CD4+ T lymphocytes, suggesting that these cells, when affected by LOY, might convey an increased risk of cancer, and this finding motivated the current study [9].

Since 2014, LOY has been investigated in the blood of aging men, but essentially nothing is known about the status of this very common mutation in leukocytes that infiltrate the tumor micro-environment (TME). TME is a complex milieu that includes different types of immune cells, including tumor infiltrating lymphocytes (TILs) and myeloid cells, but also connective tissue cells, like fibroblasts, blood vessels and tumor cells. TME can be divided into two main subtypes: ***i)*** ‘infiltrated-excluded’, known as poorly immunogenic, with CD8+ cytotoxic T lymphocytes (CTLs) located mainly along the tumor margin; and

***ii)*** ‘infiltrated-inflamed’, with highly activated CTLs, expressing PD-1 IC receptor [20]. As reported previously for CRC patients, the TME is predominantly infiltrated with circulating regulatory T cells (Tregs) [21, 22] that inhibit the activity of CD4+ T helper cells (Th) and CTLs [23, 24]. It was observed that increased number of Tregs in TME may show a dual outcome for CRC patients, either with improved [25] or worsened [26] prognosis. This could be related to differences in the expression level of IC receptors on TILs [27], which under normal conditions, induce the inhibitory signals for effector T cells. Therefore, by blocking ICs with monoclonal antibodies, the antitumor immune response of T cells can be enhanced [28, 29]. Unfortunately, not all patients benefit equally from the treatment [30]. As proposed by Kumagaj et al., 2020, the successful therapy using IC inhibitors with anti-PD-1 antibodies can be predicted by the balance between the PD-1 expression levels on CTLs and Tregs in TME [31]. In this context, revealing new factors influencing the normal function of Tregs and consequently treatment response, could improve the effectiveness of the targeted immunotherapy.

The current study addresses the gap in knowledge about the LOY status of immune cells that infiltrate TME. We show that leukocytes with LOY can be found in the primary tumors and non-tumorous uninvolved margin tissue (UM) of CRC patients as well as LM_CRCs. We performed an integrative computational and experimental study showing that the lymphocyte subset that is the most affected by LOY in tumor, UM as well as in blood are Tregs. Our meta-analysis of two single-cell RNA sequencing (scRNA-seq) datasets focused on the analysis of infiltrating leukocytes in TME. Since LOY causes dysregulation of immune genes [9, 13, 15, 16], we hypothesized that it may also affect expression level of markers in TILs related to the efficacy of immunotherapy treatment.

## Results

### Study design for CRC patients

Aiming to establish the presence and functional effects in LOY on leukocytes that infiltrate solid cancer tissue, we performed computational and experimental analysis of multiple tissues: primary or metastatic tumor, non-tumorous uninvolved margin tissue (UM) surrounding tumor in colon or liver as well as blood (Fig. 1). We re-analyzed two publicly available scRNA-seq datasets: ***i)*** leukocytes obtained from different tissues of CRC patients [32]; and ***ii)*** tumor-infiltrating immune cell atlas derived from various types of cancer [33]. We applied to both datasets a previously developed method for LOY-scoring based on the lack of expression of genes located in the male-specific region of chromosome Y (MSY), as the LOY-signature in a single cell [9, 11, 13, 34]. Using these datasets, we also investigated the transcriptomic profile of selected genes in LOY vs. non-LOY-cells in TME.

**Fig. 1.**
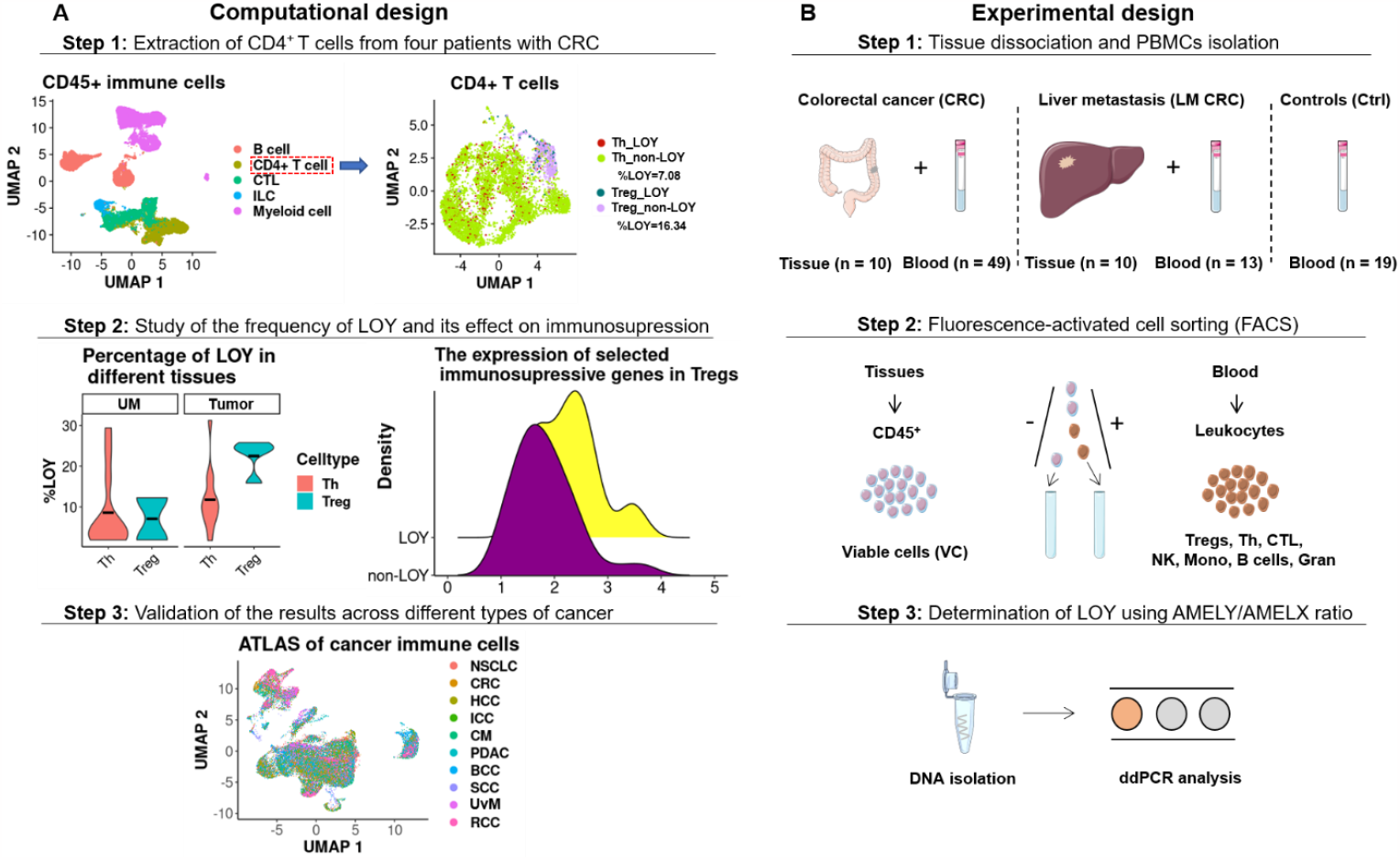
Schematic workflow of computational (Panel A) and experimental (Panel B) design applied in our study. **Panel A:** Computational analysis of two scRNA-seq datasets; Zhang et al., 2020 and Nieto et al., 2021. In the first step, CD4+ T cells from the tumor microenvironment (TME), uninvolved margin (UM) and blood samples obtained from four CRC patients (Zhang et al., 2020) were re-clustered and categorized into Th and Tregs. Next, LOY-cells were defined based on the lack of expression of MSY-specific genes. Subsequently, the status of selected immunosuppressive genes was investigated in LOY and non-LOY Tregs located in the UM and tumor sites. The results obtained from the Zhang et al., 2020 dataset and comprising the scRNA-seq were validated in the Nieto et al., 2021 dataset, the latter covering an atlas of CD45+ immune cells from eight types of cancer, including CRC. **Panel B:** Experimental design for analysis of cells obtained from TME of human CRCs (n = 10) and LM_CRCs (n = 10) tissues and blood samples collected from CRC (n = 49), LM_CRC (n = 13) and Ctrl (n = 19) patients showed in three consecutive steps. Suspensions of single cells were sorted by FACS, followed by DNA isolation, ddPCR analysis using TaqMan-probes. Parts of the figure were drawn by using pictures from Servier Medical Art. Servier Medical Art by Servier is licensed under a Creative Commons Attribution 3.0 Unported License (https://creativecommons.org/licenses/by/3.0/). Abbreviations: TME – tumor microenvironment; UM – non-tumorous uninvolved margin tissue; CRC – colorectal cancer; LM_CRC – liver metastasis of CRC; Ctrl – control patients, Tregs – regulatory T cells; Th – helper T cells; CTL – CD8+ cytotoxic T cells; ddPCR – droplet digital PCR; scRNA-seq – single-cell RNA-seq.

Moreover, we extended the analyses by performing tissue dissociations on tumors and UM fragments from two independent sets of 10 primary CRC- and 10 LM_CRC patients. Additionally, we implemented a protocol, which enabled the isolation of CD45+ immune cells obtained from both tumors and UMs as well as sorting of seven subpopulations of leukocytes from blood (Fig. 1). The analysis of LOY status was performed using previously described droplet-digital PCR (ddPCR) method [12, 13]. Blood samples for sorting experiments were collected from 49 CRCs, 13 LM_CRCs and 19 control patients. All study subjects were qualified for our research according to established inclusion criteria (Materials and Methods, Table 1, Table S1). The variable level of leukocytes was confirmed in dedicated histopathological analysis of the hematoxylin/eosin-stained sections from tumor and UM (Table 1). Summary of clinical characteristics for all cancer patients and controls can be found in Table S1.

**Table 1.** Summary of clinical data and experimental results from 10 CRC and 10 LM_CRC patients qualified for the dissociation experiments.

| Patient ID | Diagnosis | Smoking | ICD10 | %Immune Infiltrates<br>[Histpat H/E] | Blood<br>[ddPCR %LOY] | UM<br>[ddPCR %LOY] | Tumor<br>[ddPCR %LOY] |
| --- | --- | --- | --- | --- | --- | --- | --- |
| 1 | CRC | Yes | C20 | 10 | 0 | 0 | 0 |
| 2 | CRC | No | C18.2 | 5 | 0 | 0 | 0 |
| 3 | CRC | No | C18.3 | 10 | 0 | 0 | 0 |
| 4 | CRC | Yes | C18.7 | 5 | 8 | 0 | 0 |
| 5 | CRC | Yes | C18.7 | 1 | 11 | 7 | 6 |
| 6 | CRC | Yes | C20 | 3 | 10 | 11 | 11 |
| 7 | CRC | Yes | C18.0 | 10 | 13 | 0 | 11 |
| 8 | CRC | Yes | C18.9 | 10 | 6 | 6 | 16 |
| 9 | CRC | Yes | C18.2 | 3 | 10 | 0 | 0 |
| 10 | CRC | Yes | C20 | 5 | 11 | 15 | 13 |
| 11 | LM CRC | Yes | C78.7 | 5 | 0 | 0 | 0 |
| 12 | LM CRC | No | C78.7 | 10 | 6 | 0 | 8 |
| 13 | LM CRC | Yes | C78.7 | 3 | 0 | 0 | 0 |
| 14 | LM CRC | Yes | C78.7 | 10 | 11 | 14 | 17 |
| 15 | LM CRC | Yes | C78.7 | 15 | 8 | 0 | 0 |
| 16 | LM CRC | Yes | C78.7 | 15 | 9 | 0 | 0 |
| 17 | LM CRC | No | C78.7 | 0 | 10 | 10 | 0 |
| 18 | LM CRC | No | C78.7 | 5 | 9 | 0 | 30 |
| 19 | LM CRC | No | C78.7 | 5 | 0 | 0 | 0 |
| 20 | LM CRC | Yes | C78.7 | 20 | 44 | 45 | 23 |

ICD10 – International Statistical Classification of Diseases and Related Health Problems. Immune infiltrates were assessed by the pathologist during histopathological analysis in the hematoxylin/eosin-stained sections from tumor and surrounding normal tissue. UM – uninvolved margin of normal tissues surrounding tumor. Diagnoses: CRC – colorectal cancer, LM\_CRC – liver metastasis of CRC.

### Tregs showed the highest percentage of LOY-cells among CD4+ T cells in primary CRCs

As previously reported, higher levels of LOY were found in the population of CD4+ T lymphocytes (CD4+ T cells) among all leukocytes from men with prostate cancer, versus controls [9]. Therefore, we performed the analysis of scRNA-seq dataset ([32], see Materials and Methods) to determine the distribution of LOY-cells in the three most common subpopulations of leukocytes infiltrating the tumors, namely Tregs, Th and CTLs. We focused on four male patients from the Zhang-dataset [32] and two groups of previously annotated cells, i.e. CD4+ T cells and CTLs. CD4+ T cells were reclustered and then categorized as Th and Tregs. Tregs were distinguished from the group of Th cells based on the expression of *CTLA4* and *IL23R* markers (Table S2). All analyzed groups of cells were further divided into two groups based on the lack of expression of six genes located in the MSY and indicating LOY status (see Materials and Methods). As detailed in Table S2, Tregs were represented by a pooled dataset of 498 cells collected from four patients, whereas Th and CTLs were represented by 1460 cells from four CRC patients and 681 cells from three patients, respectively. Interestingly, we found that Tregs in the tumors had a higher level of LOY-cells, in comparison to Th, with the mean level of LOY of 22% for Tregs and 11% for Th cells (p=0.033, Wilcoxon test with Bonferroni correction). In the case of CTLs, the mean LOY value was 14% and the difference between Tregs and CTLs showed similar statistical significance (p=0.023, Fig. 2).

**Fig. 2.**
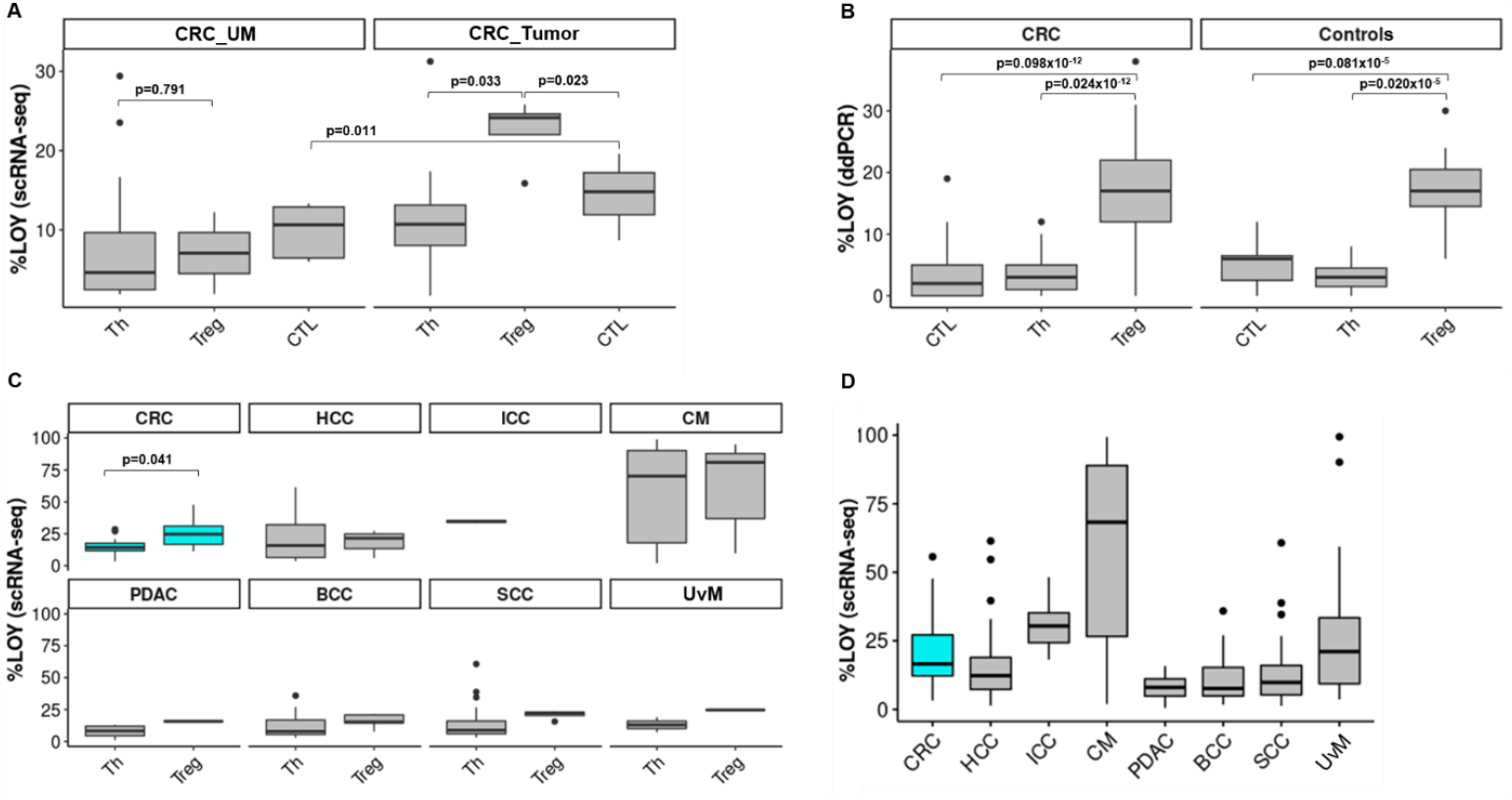
Experimental and computational analysis of distribution of LOY in subpopulations of T lymphocytes. **Panel A:** Meta-analysis of the distribution of LOY in Tregs (n = 2, cells = 203), Th (n = 4, cells = 1818) and CTL (n = 3, cells = 1441) lymphocytes located in the uninvolved margin (UM) and Tregs (n = 4, cells = 498), Th (n = 4, cells = 1460) and CTL (n = 3, cells = 681) located in tumor of male patients with CRC. **Panel B:** Experimental analysis of the distribution of LOY in Regulatory T lymphocytes (Tregs), Helper T lymphocytes (Th) and CD8+ cytotoxic T cells (CTL) in blood samples collected from 49 CRC patients and 19 healthy controls. CRC – colorectal cancer, Ctrl – age-matched control patients without prior diagnoses of cancer and Alzheimer’s disease. **Panel C:** Meta-analysis of distribution of LOY in Tregs and Th in eight different cancer types: CRC -colorectal cancer (n = 10, cells = 3349); HCC – hepatocellular carcinoma (n = 9, cells = 8329); ICC – intrahepatic cholangiocarcinoma (n = 1, cells = 222); CM – cutaneous melanoma (n = 24, cells = 13306); PDAC – pancreatic ductal adenocarcinoma (n = 4, cells = 845); BCC – basal cell carcinoma (n = 8, cells = 14053); SCC – squamous-cell carcinomas (n = 8, cells = 13889); UvM – uveal melanoma (n = 1, cells = 248). **Panel D:** Meta-analysis of distribution of %LOY in CD45+ immune cells located in TME of different cancers. CRC – colorectal cancer (n = 10, cells = 11165), HCC – hepatocellular carcinoma (n = 16, cells = 21604), ICC – intrahepatic cholangiocarcinoma (n = 5, cells = 1497), CM – cutaneous melanoma (n = 33, cells = 41087), PDAC – pancreatic ductal adenocarcinoma (n = 11, cells = 4749), BCC – basal cell carcinoma (n = 8, cells = 35359), SCC – squamous-cell carcinomas (n = 4, cells = 25924), UvM – uveal melanoma (n = 4, cells = 5018).

Similarly, we studied the UMs of CRC patients. We did not observe similar differences in the distribution of LOY in Tregs, Th or CTLs in UMs, as in the tumors (Figs. 2 and 3). In general, we found a low level of LOY in all analyzed cell types of the UMs, where Tregs were represented by a pooled dataset of 203 cells. However, this result is based on only two patients. Th- and CTL-cells from UMs were represented by 1818 pooled cells from four CRC patients and 1441 pooled cells from three patients, respectively.

**Fig. 3.**
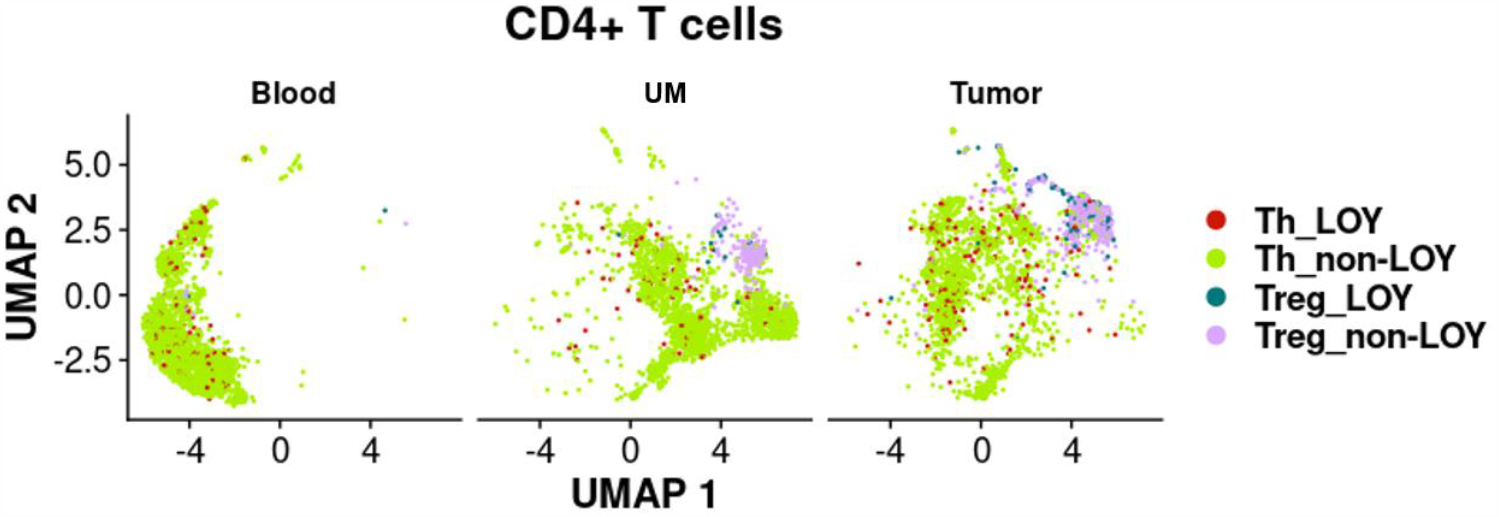
Computational analysis of distribution of LOY CD4+ T lymphocytes in different tissues of CRC patients. CD4+ T cells were extracted from the tumor, uninvolved margin (UM) and blood samples obtained from four CRC patients (Zhang et al., 2020) and categorized into Th and Tregs. Next, LOY-cells were defined based on the lack of expression of MSY-specific genes.

### Tissue dissociations confirm the presence of LOY among CD45+ immune cells from primary tumors and liver metastases of CRC

As mentioned above, LOY has been associated with different types of cancers, but the presence of LOY in leukocytes infiltrating solid tumors was not studied. We aimed here to validate the computational analysis of public scRNA-seq datasets by examination of LOY in CD45+ immune cells from primary tumors, UMs and whole blood as well as in sorted leukocytes isolated from matched peripheral blood of CRC patients. Since CRC patients have ∼50% risk of liver metastases, we also enrolled patients with diagnosed LMs (Table 1). Out of 20 patients, five with CRCs and four with LM_CRCs showed detectable LOY in CD45+ immune cells infiltrating the tumors using ddPCR, with the mean percentage of LOY (%LOY) in the range of 6-30% (Table 1). However, the remaining 11 cases did not reveal LOY above the general detection threshold of 5% measured with ddPCR, suggesting a heterogeneity of CRCs/LM_CRCs, when %LOY-cells that infiltrate the solid tumors is considered. Corresponding analysis of CD45+ immune cells infiltrating UMs from the same set of patients showed seven cases with detectable LOY. The level of LOY-cells in UMs was generally lower (6-15%) than in tumors, with the notable exception of UM from patient ID 20 (45% of LOY-cells) (Table 1). Analysis of bulk DNA from whole blood of the same patients also showed seven patients with LOY, and there were no differences in the %LOY between blood DNA and CD45+ immune cells infiltrating the tumors. As could be expected from the above results, there were positive correlations between LOY in CD45+ immune cell infiltrates of the tumors and UMs (n=10, r=0.71, p=0.02) as well as between immune cell infiltrates in the tumor and blood DNA (Fig. S1).

We supported the above results by additional comprehensive analysis of LOY in leukocytes of CRC patients and controls (free of cancer and Alzheimer’s disease) aiming at analysis of sorted Tregs along with other cellular subsets. We analyzed the distribution of LOY in the peripheral blood of 49 CRC patients and 19 healthy individuals in seven sorted subsets of cells, as well as in the bulk DNA from blood. The LOY analysis using ddPCR revealed that Tregs had significantly higher %LOY in comparison to Th- and CTL-cells in both CRC patients and controls (p=0.024×10^−12^ and p=0.02×10^−5^, respectively) (Fig. 2B). The %LOY for Tregs in CRC patients was 17% and 4% for the Th-cells. Similar comparison in case of controls showed the level LOY for Tregs was 17% and 3% for Th-cell fraction. In another comparison Tregs with CTLs, the mean level of LOY for CRC patients was 4% and for controls 5% (p=0.098×10^−12^ and p=0.081×10^−5^, respectively) (Fig. 2B). Tregs showed an overall higher %LOY in blood in comparison to all other sorted cell subpopulations. In a similar fashion, we investigated blood samples derived from 13 LM_CRC patients (Fig. 4). In conclusion, our experimental data suggest that Tregs have a significantly higher %LOY-cells in blood in comparison to Th and CTLs in CRC and LM_CRC as well as control patients. The similar levels of LOY in Tregs from CRC patients and controls is, however, a surprising finding.

**Fig. 4.**
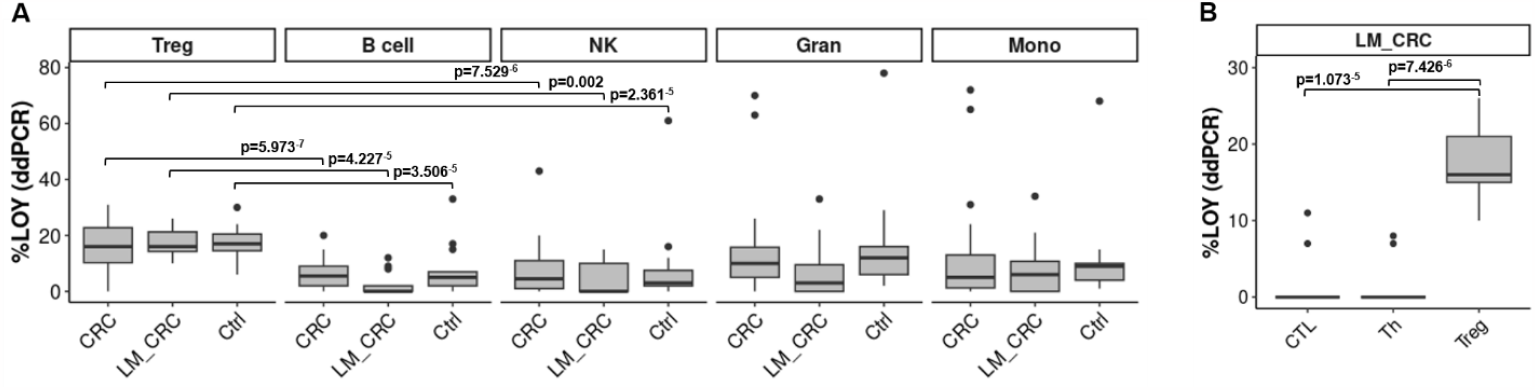
Experimental detection of the distribution of LOY in sorted blood cells from patients with primary CRC, liver metastases of CRC and controls. **Panel A: Distribution of LOY in five subpopulations of CD45+ blood leukocytes**. LOY was analyzed in sorted B cells, Gran, Mono, NK, Tregs obtained from 49 male CRC patients, 13 LM_CRC patients and 19 healthy controls. CRC – colorectal cancer, LM_CRC – liver metastasis of CRC, Ctrl – control patients. Pairwise comparison (Wilcoxon test with Bonferroni correction) is shown only for Tregs vs. B cells and Tregs vs. NK cells. These subsets (B cells and NK cells) displayed the highest statistical significance upon comparison with Tregs. **Panel B: Distribution of LOY in FACS sorted subpopulations of T lymphocytes**. Analysis of the distribution of LOY in Tregs, Th and CTLs in blood samples collected from 13 LM_CRC patients.

### Tregs with LOY show higher expression of immunosuppressive genes in TME than non-LOY Tregs in CRC patients

To identify the functional effect of LOY on Tregs in the TME, we re-analyzed the dataset of Zhang et al., 2020 [32] to reveal changes between the expression levels of genes related to LOY Tregs vs. non-LOY Tregs (see Materials and Methods). First, we separated 101 LOY Tregs from 397 non-LOY Tregs in four male CRC patients. We focused in particular on genes related to the suppressive features of Tregs, among others, genes encoding IC receptors (*CTLA4, PDCD1, HAVCR2, LAG3, TIGIT* and *ICOS*), genes connected with manipulation of antigens presenting cells (*ENTPD1, NT5E, CD80*) and migration abilities (*CCR1, CCR4, CCR5, CCR7, CCR8*). We also studied the expression level of *IKZF2* gene, the member of Ikaros family of zinc-finger proteins, which encodes a transcription factor named Helios. Based on the gene expression score (Material and Methods), we focused on the three genes, *PDCD1, TIGIT* and *IKZF2*, encoding PD-1 and TIGIT surface markers and Helios transcription factor, respectively. *TIGIT* and *IKZF2* showed a significantly higher expression in LOY Tregs than non-LOY Tregs (p=0.004 and p=0.031 respectively) (Fig. 5). For the *PDCD1* we observed a similar trend but our results were not significant (p=0.069). PD-1 and TIGIT are known immune checkpoint inhibitors characteristic for suppressive phenotype of Tregs in TME. Also, co-expression of PD-1 with Helios^high^ molecules is characteristic for suppressive features of Tregs in TME.

**Fig. 5.**
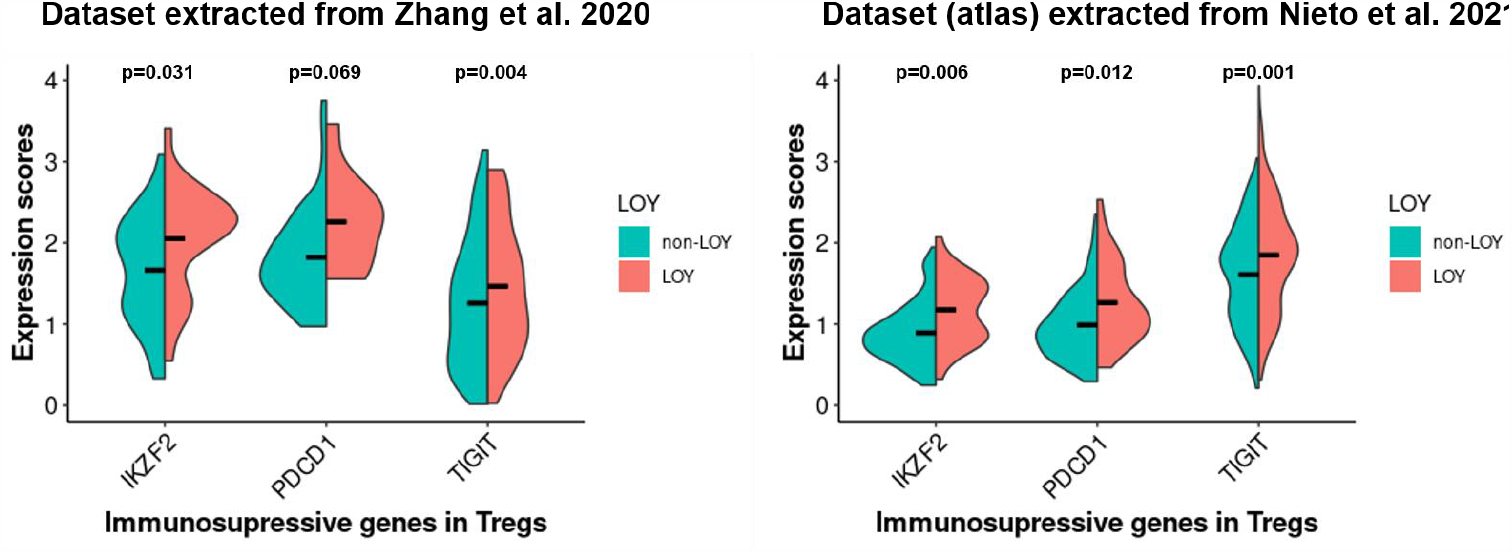
Estimation of the level of immunosuppressive gene expression in LOY vs non-LOY Tregs located in TME of CRC patients. Horizontal lines in violin plots show mean values. Left panel represents the data extracted from Zhang et al., 2020 in the tumors of four CRC patients (LOY cells = 101; non-LOY cells = 397). Right panel is the validation of the results with the data obtained from Nieto et al., 2021, representing data extracted from the tumors of 11 CRC patients (LOY cells = 396; non-LOY cells = 887).

A corresponding analysis of the scRNA-seq dataset extracted from Nieto et al., 2021 [33] confirmed the above observations about significantly higher expression of *PDCD1, TIGIT* and *IKZF2* in LOY Tregs located in TME (Fig. 5). In this atlas dataset, we validated our results on a group of 11 CRC patients, with 396 LOY Tregs and 887 non-LOY Tregs. Our study also revealed four additional genes (*IL32, CD27, MAGEH1* and *LAYN*) that were the top-ranked genes correlated with the expression of *TIGIT* in Tregs located in the TME (Table S3). These genes were reported previously to be associated with the regulation of Tregs function and induction of their suppressive phenotype followed by a poor prognosis of CRC patients [35]. In conclusion, the results suggest that LOY may intensify features related to the suppressive phenotype of Tregs in TME of CRC patients.

### Tregs in TME from CRC and melanoma display highest levels of LOY

We further explored the dataset from the tumor immune cell atlas [33] for the presence of LOY in CD45+ immune cells, and Tregs in particular, within TME of CRC and seven other types of cancer (Fig. 2C). These other diagnoses were hepatocellular carcinoma (HCC); intrahepatic cholangiocarcinoma (ICC); cutaneous melanoma (CM); pancreatic ductal adenocarcinoma (PDAC); basal cell carcinoma (BCC); squamous-cell carcinoma (SCC); and uveal melanoma (UvM). LOY was scored as described above in 146,403 of CD45+ immune cells. With this analysis, we verified the presence of LOY in immune cells within TME in eight cancer diagnoses (Fig. 2C). The CRC diagnosis was represented by a pooled dataset of 3349 cells from 10 patients (Table S4). In agreement with our approach described above, we tested whether Tregs had a significantly higher percentage of cells with LOY in comparison to Th also in eight types of cancer. Interestingly, a significant difference between the percentage of cells with LOY in Tregs and Th was found only in the CRC diagnosis (p=0.041) (Fig. 2C). Combined with other data, the results suggest that Tregs with a significantly higher percentage of cells with LOY in comparison to Th-subset might be a part of characteristic signature of CRC diagnosis.

## Discussion

The findings of LOY in Tregs from peripheral blood and TME of CRC patients considerably expand our previous results concerning the higher levels of LOY in the CD4+ T cells in men with prostate cancer vs. controls [9]. We might explain here why LOY in CD4+ T cells is associated with increased risk of cancer, although CD4+ T cells in these patients are affected by LOY at relatively low frequency, compared to other subsets of leukocytes [9]. Tregs represent 5-10% of CD4+ T cells in blood of healthy subjects [36], but in cancer patients, the frequency of circulating Tregs increases by approximately 2-fold [37, 38]. Furthermore, increased levels of Tregs in TME has been positively correlated with poor prognosis and low survival rates in various cancers [36, 39-41]. However, these studies were performed in cohorts combining men and women and LOY was not examined. Hence this aspect requires further investigation.

The importance of TME in the improvement of CRC treatment is emerging and Tregs are an attractive therapeutic target in cancer immunotherapy [35, 36]. Depending on the interaction with factors in the local environment, Tregs in TME can also differentiate into distinct subpopulations of naïve, effector and memory cells. Tumor-associated Tregs are known to be predominantly composed of effector cells, which upregulate surface molecules associated with enhanced immuno-suppressive activity. This can lead to a worse immunological response followed by facilitation of tumor growth [42]. On the other hand, effector Tregs are also known for IL-10 secretion in TME of CRC patients, resulting in the inhibition of the Th17 cells, which are responsible for the inflammation and tumor immunity [43]. It should be stressed that in our analysis CRCs had a higher number of Tregs carrying LOY in TME, comparing to the rest of the CD4+ T lymphocytes.

The blockade of PD-1 immune checkpoint receptor with monoclonal antibodies can lead to enhanced anti-tumor immune response [28, 29] and therefore immunotherapy became an attractive alternative treatment method. Unfortunately, many patients (both males and females) do not respond properly to immunotherapy and the reason(s) for this is not clear, but in the case of CRC, it may be related to the phenotype of infiltrating Tregs [28]. We showed here that LOY affects expression levels of *PDCD1, TIGIT* and *IKZF2* genes in Tregs. Two of these genes (*PDCD1* and *TIGIT*), which are IC receptors and together with transcription factor *IKZF2* are involved in the formation of suppressive phenotype of Tregs in TME and this presumable effect of LOY might be related to the efficacy of immunotherapy.

In our analysis, the 19 control subjects that were free from diagnoses of cancer and Alzheimer’s disease have essentially the same LOY-levels in circulating Tregs as cancer patients, which is a surprising finding (Fig. 2B). Multiple explanations could be proposed here: ***i)*** The strongest factor associated with increasing levels of LOY in blood is age and the mean age of our controls is higher than cancer patients (see Table S1). However, it is unlikely that it is the sole explanation for this result; ***ii)*** Our control cohort is relatively small and this issue should be followed up in a considerably larger cohort of CRC patients and controls; ***iii)*** The relative number of cells in each category of tumor infiltrating cells (Tregs, Ths, CTLs) should also be taken into account, as it has been suggested that cancer patients have higher number of Tregs, compared to healthy subjects (see above); ***iv)*** Tumor immune-surveillance is a defensive mechanism (and not driving the tumor development), which might not function properly in some healthy subjects, but in the absence of the processes that drive the tumorigenesis, there might be no negative consequences (i.e. tumors) for such a subject.

It should also be mentioned that CTLs showed an increased level of LOY in TME of CRC patients (Fig. 2A) compared to UM (p=0.011) and CTLs are one of the critical components of tumor immune-surveillance within TME. CTLs, upon stimulation with an antigen, are undergoing clonal expansion and this might be related to their elevated level of LOY, since LOY is presumed to occur as a result of an error during mitotic division. In parallel, LOY might compromise some of the normal functions of CTLs within TME and this aspect deserves future studies using scRNA-seq exploring T-cell receptor (TCR) sequencing combined with global transcriptome analysis. We have shown previously that circulating CTLs in blood had very low levels of LOY in prostate cancer patients, Alzheimer’s disease patients and age-matched healthy controls [9], but the situation might be different within TME.

Loss of chromosome Y has also been shown as a very frequent mutation in tumor cells from numerous cancers and LOY correlates with an overall worse prognosis [44, 45]. This agrees with the concept that LOY provides proliferative advantage to cells affected by this aneuploidy [46]. This is also in line with the fact that men have a higher incidence and/or mortality from most sex-unspecific cancers, which is unexplained by known risk factors [47, 48]. This sex disparity is an understudied aspect, which might be related to LOY occurring both in the tumor cells and in cells responsible for defensive mechanism of tumor immune-surveillance within TME. Thus, future studies should pay attention to the analysis of LOY in tumor cells and in the normal immune cells infiltrating TME. It is interesting in this context that our meta-analysis of Nieto et al. dataset of multiple cancers [33] showed that, in addition to CRC, melanoma display high levels of LOY, which should stimulate future analyses. It is noteworthy that cutaneous malignant melanoma display sex bias. By the age of 65 and by the age 80, men are twice and three times more likely to get melanoma, respectively, when compared to women. Moreover, males of any age have also a greater risk of death from this disease [49].

We performed an integrative analysis of LOY-cells in tumor, normal tissue (unchanged large bowel mucosa) surrounding tumor and blood, using experimental tissue dissociation and publicly available scRNA-seq datasets, aiming to identify and characterize LOY-affected leukocytes. Experiments using scRNA-seq methods has recently revolutionized the medical field [50] and carry a promise of extending the research into personalized diagnostics [34]. scRNA-seq allows to overcome limitations regarding the investigation of the LOY frequency and LOY-associated transcriptomic effects, especially when the number of cells is not sufficient for the bulk analysis, as is often the case of immune cells isolated from solid tissues, or when the data from homogenous cell populations is required [13]. On the other hand, the advantage of using ddPCR is its simplicity and a low input of DNA, allowing detection of LOY in DNA from sorted cells, starting at about 10,000 cells [13]. Results presented here and during the past few years regarding chromosome Y showed that it should no longer be considered as a “genetic wasteland” [9].

In conclusion, our study sets the direction for further research regarding a probable functional role of LOY in intensifying features related to the suppressive phenotype of Tregs in TME and, as a consequence, a possible influence on immunotherapy response in CRC patients [42]. The presence of LOY has previously been associated with a considerable dysregulation of autosomal gene expression in leukocytes, in particular responsible for the immune-surveillance functions of circulating cells [9, 15, 16]. Here, we extend these finding to TME of CRC patients and this aspect warrant further studies.

## Materials and methods

### Human specimens

For the tissue dissociation, 20 male patients were enrolled including 10 patients with a primary CRCs recruited in the Oncology Center in Bydgoszcz and 10 patients with metachronous (seven patients) and synchronous (three patients) liver metastases of CRCs (LM_CRC) recruited in the University Clinical Center in Gdańsk. Seven metachronous LM_CRC patients were treated with an adjuvant chemotherapy before the surgery; four LM_CRC individuals were diagnosed with rectum adenocarcinoma and six with colon adenocarcinoma. The inclusion criteria for the recruitment of CRC was tumor size of at least 3 cm in diameter. The neoadjuvant therapy was considered as an exclusion criterion for primary CRCs. The age of patients with primary CRC ranged from 42 to 74, with a median age of 62, and from 42 to 78, with a median age of 65 for LM_CRC group. Detailed clinical information is shown in Tables 1 and S1. Additionally, blood samples were collected from 49 CRC, 13 LM_CRC and 19 control (Ctrl) patients recruited in the University Clinical Center in Gdańsk. Ctrl patients were collected from the general male population of Gdańsk and their age ranged from 62 to 74, with a median age of 68, as described [51]. Written informed consent was obtained from all the patients prior to material collection. Our study followed principles of the Declaration of Helsinki and was approved by the Independent Bioethics Committee for Research at the Medical University of Gdańsk (approval number NKBBN/564/2018 with multiple amendments).

For tissue dissociation we used two fragments excised from the same patients: UMs located at least 3-5 cm away (edge to edge distance) from the tumor and a fragment of primary tumor. Each collected sample was divided into two parts: one for tissue dissociation, one for FFPE block. The splitting surface underwent histological assessment of the actual tissue content and immune cells infiltration. This approach provides the correct histopathological assessment of the actual content of our material. The histopathological assessment was carried out by experienced pathologists. The absence of tumor cells was confirmed in all UM fragments. Samples for dissociation were collected directly in the MACS Tissue Storage Solution (Miltenyi Biotec), transported on ice and, according to the manufacturer’s instructions, processed within 24 hours.

### Tissue dissociation

Samples for tissue dissociation studies were washed with phosphate-buffered solution (PBS), minced into one mm fragments on ice and mechanically dissociated using gentleMACS Dissociator (Miltenyi Biotec). Enzymatic digestion was performed using the Tissue Dissociation Kit (Miltenyi Biotec) at 37°C with agitation. Digestion process was stopped after two 30 min incubations in RPMI-1640 (Sigma Aldrich) with 10% FBS (Sigma Aldrich). The cell suspension was filtered with 70 µM nylon cell strainer (BioLogix) and centrifuged at 300 x g in 4°C for 7 min. Red blood cells were lysed using one ml of lysing solution (BD Biosciences).

### FACS sorting of tissue infiltrating leukocytes

Cell suspensions obtained from tumor and UM fragments after dissociation were incubated with a Fixable Viability Dye eFluor^tm^ 780 (Invitrogen, ThermoFisher Scientific) to label dead cells and with anti-CD45+ antibodies (BD Biosciences) to distinguish leukocyte populations. Cells were incubated with the dye and antibody for 30 min at 4°C, washed and sorted to PBS with 0.04% BSA (Sigma Aldrich), centrifuged at 3200 x g in 4°C for 10 min and stored in -80°C for DNA extraction.

### FACS sorting of peripheral blood leukocytes

Granulocytes and peripheral blood mononuclear cells (PBMCs) were collected from 6 ml and 32 ml blood into EDTA-K2 tubes (BD Biosciences) and BD Vacutainer^®^ CPT™ Mononuclear Cell Preparation Tubes (BD Biosciences), respectively. A sample of 1.5 ml whole blood (EDTA-K2 tubes) was taken for DNA extraction. The number of PBMCs obtained by density gradient centrifugation method was estimated using EVE Automatic Cell Counter and immediately processed for FACS. The number of 0.5×10^6^ PBMCs were centrifuged at 3200 x g in 4°C for 10 min and stored in -80°C for DNA extraction. The population of CD4+ T cells was isolated using negative immunomagnetic separation KIT (Miltenyi Biotec) and incubated for 30 min at 4°C with Human Regulatory T Cell Cocktail (BD Biosciences). Positively separated cells were labeled with antibodies for Monocytes (Mono); Natural killers (NK); CD19+ B lymphocytes (B cells); and CD8+ cytotoxic T lymphocytes (CTL) (BD Biosciences). Labeled cells were washed and resuspended in one ml of PBS+0.04% BSA. CD4+T cells were sorted into Treg (CD4+, CD25+, CD127^low^) and Th (CD4+, CD25+, CD127+) cell populations using Aria III analyzer (Beckton Dickinson). The remaining PBMCs were sorted into Mono (CD45+, CD3-, CD14+), B cells (CD45+, CD3-, CD19+), CTL (CD45+, CD3+, CD8+) and NKs (CD45+, CD3-, CD56+, CD16+). Granulocytes were sorted according to SSC/FSC parameters and CD45+ expression antigen (BD Bioscience).

### Measurement of LOY using ddPCR

Quantification of the level of LOY in analyzed samples was performed using previously described procedure [12]. Briefly, DNA extracted from sorted fractions of cells was digested with *Hind*III enzyme (#FD0504, ThermoFisher Scientific) for 15 min in 37°C. Next, 50 ng of digested and diluted DNA was thoroughly mixed with ddPCR supermix for probes no dUTP (Bio-Rad), PCR primers and probes targeting 6 bp difference between the AMELX and AMELY gene assay (C_990000001_10, ThermoFisher Scientific). For the processing of samples and fluorescent measurement of the signal in channels FAM and VIC, we used the QX200 Droplet Digital PCR System (Bio-Rad). For the generation and analysis of the data we used the dedicated software QuantaSoft (version 1.7.4.0917).

### scRNA-seq data collection and analysis of LOY

For our meta-analysis we selected two scRNA-seq datasets. The first one was obtained from Zhang et al. [32] generated by the 10x Genomics platform. The gene expression profiles made by 10x Genomics platform contained the information for 18 colorectal cancer patients, including 14 females and four males. In our study we focused only on male patients, and two groups of previously annotated cells i.e., CD4+ T and CTL cells. Further on, these two groups of cells were distinguished based on the patients, tissue of origin i.e., tumor, UM, blood and cell subpopulations. Later, groups with the sum of LOY and non-LOY-cells lower than 15 were removed from our study.

The second dataset that was used for the validation of our results was obtained from Nieto et al. [33]. It contained the gene expression profiles of 500,000 CD45+ immune cells acquired from 217 patients and 13 cancer types. Patients were separated to male, female and unknown genders. To distinguish the unknown group to male and female, we calculated the percentage of gene expression for the set of chromosome Y specific genes i.e., *RPS4Y1*; *ZFY*; *USP9Y*; *DDX3Y*; *KDM5D*; and *EIF1AY* using the default parameters of the ‘PercentageFeatureSet’ function of Seurat R package (version 4.2.0) [52]. Patients with and without expression of these six genes were defined as male and female, respectively. After removal of female patients and cancers with less than four patients, we extracted information from 98 male patients and eight cancers. According to the percentage of gene expression defined by the ‘PercentageFeatureSet’ function of Seurat R package in both datasets, the cells showing zero expression for all six chromosome Y specific genes were classified as LOY and the rest were defined as non-LOY. In this dataset, cells were distinguished based on patients, cell subpopulations and cancer types. Since the second dataset consisted of higher number of patients and cells in comparison with the first dataset, we used stricter threshold for data filtering. Hence, groups with the sum of LOY and non-LOY-cells below 50 were removed from our study.

### Immunosuppressive gene expression scores in Tregs with/without LOY

Tregs located in the tumors were distinguished from the group of CD4+ T cells based on the expression of *CTLA-4* and *IL23R* markers for both datasets used for the meta-analysis [32, 33]. Next, Tregs were divided into two groups based on the LOY status and their expression profiles were compared, as described above. We computed the gene expression scores per cell for *IKZF2, PDCD1* and *TIGIT* genes using the ‘AddModuleScore’ function of Seurat R package by default parameters. Expression scores were the average expression levels of desired genes in each cluster (i.e., LOY and non-LOY Tregs) subtracted by the aggregated expression of randomly selected control feature sets.

### Statistical analysis

We compared the percentage of LOY between Tregs and other cells in Figs. 2 and 4 in addition to gene expression scores for immunosuppressive genes between LOY and non-LOY Tregs in Fig. 5. The significance in all pairwise comparison was computed using Wilcoxon test and p-values were adjusted using the Bonferroni correction method. For this purpose, the ‘wilcox.test’ function of R with default parameters was used.

## Data Availability

All data produced in the present study are available upon reasonable request to the authors

## Author contributions

M.W., U.J., K.D-C., K.C. conducted laboratory experiments; J.M., J.P.D. conceived of the idea; E.M., M.W., U.J. analyzed the data; J.P.D. obtained funding; M.W., U.J., J.M., N.F., J.P.D. supervised the project; M.W., U.J., E.M., J.M., J.P.D. contributed to interpretation of the results; M.W., U.J., E.M., E.R-B., B.B-O., H.D., K.C., P.O., N.F., J.P.D. drafted the manuscript; M.W., U.J., K.D-C., E.R-B., M.B., M.J., O.R., A.H., R.P., J.K., M.Z., J.K., W.Z., W.B., Ł.S., P.R., N.F. contributed to selection and collection of samples from patients; All co-authors read and approved the submission.

## Funding sources

This study was supported by grants from the Foundation for Polish Science under the International Research Agendas Program (grant number MAB/2018 /6; co-financed by the European Union under the European Regional Development Fund), Swedish Heart-Lung Foundation (grant number 20210051), Swedish Research Council (grant number 2020-02010), Swedish Cancer Society, Hjärnfonden, and Alzheimerfonden to J.P.D.

## Declaration of competing interests

J.P.D is a cofounder and shareholder in Cray Innovation AB. The remaining authors declare that they have no competing interests.

## Acknowledgements

We thank the anonymous patients and volunteer healthy controls for sample contribution and information provided in the questionnaire. We are thankful to Dr. Daniil Sarkisyan for critical review of the manuscript. We also thank physicians and nurses involved in the patient recruitment process, collaborating technicians, diagnosticians and pathologists from Oncology Center-Prof. Franciszek Łukaszczyk Memorial Hospital in Bydgoszcz (Jowita Nowaczewska, Katarzyna Krzysiak, Anetta Słupicka, Mateusz Matusiak) and University Clinical Centre in Gdańsk (Grażyna Stęplewska, Elżbieta Wierszyło, Elżbieta Pietruszka, Grażyna Dombrowska, Justyna Pietruszewska, Irena Pellowska, Michał Kunc, Aleksandra Korwat, Ewa Miłoszewska).

---

**Fig. S1.**
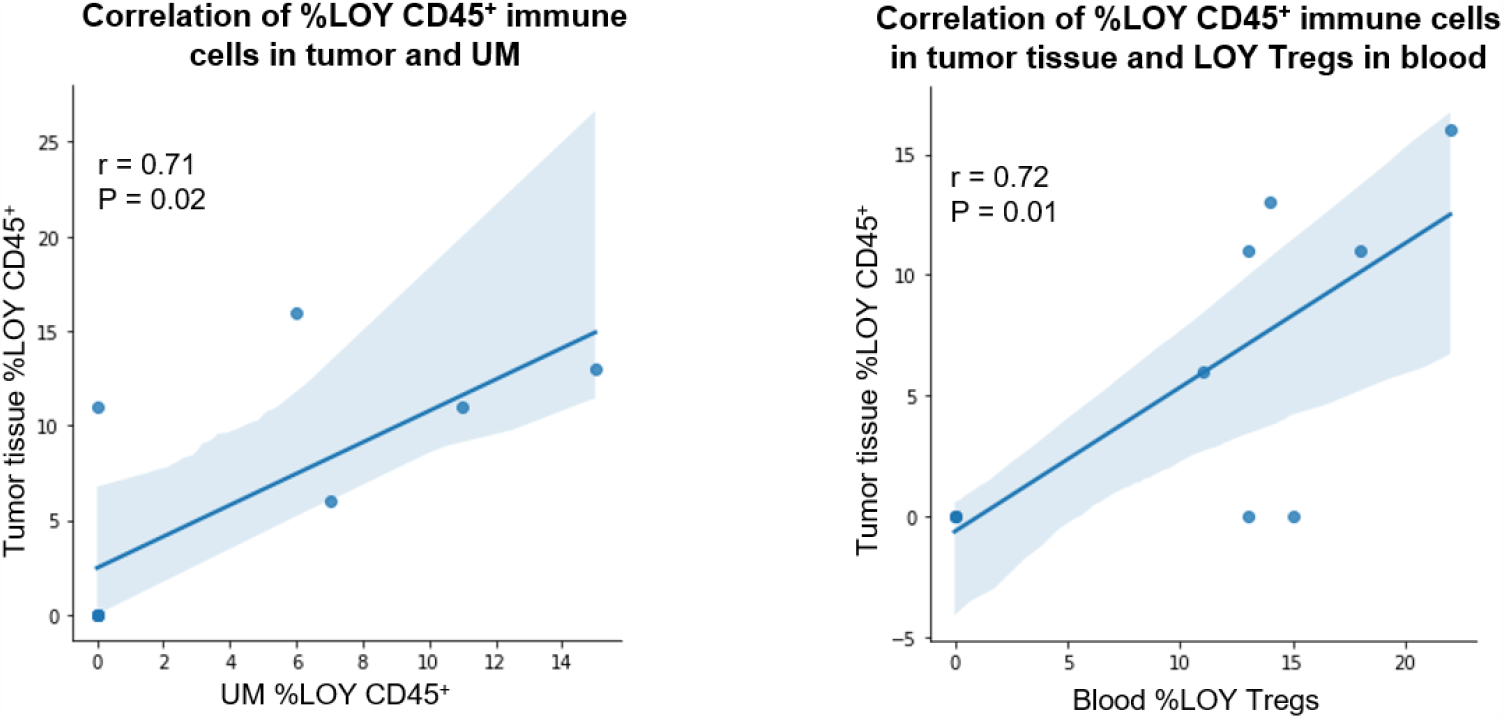
Analysis of association between the level of %LOY and cell types located in different types of tissues. Left panel represents the association between the level of %LOY detected in tumor and uninvolved margin (UM) obtained from four men diagnosed with CRC. Right panel shows the association between the level of %LOY detected in tumor and Tregs obtained from four men diagnosed with CRC. Shaded areas indicate 95% confidence intervals.

**Table S1.** Basic clinical characteristics of CRC, LM_CRC and Ctrl patients qualified for the dissociation and FACS experiments. Diagnosis: CRC – colorectal cancer, LM_CRC – liver metastasis of CRC, Ctrl – control patients.

| Diagnosis | No. of subjects | Inclusion criteria | Exclusion criteria | Average age (age range) | Type of material collected |
| --- | --- | --- | --- | --- | --- |
| CRC | 10 | Sex = male, primary tumor size $\geq 3$ cm | Neoadjuvant therapy | 62 (42 – 74) | Tissues |
| CRC | 49 | Sex = male | N/A | 67 (30 – 94) | Blood |
| LM_CRC | 10 | Sex = male, primary tumor = CRC | N/A | 65 (42 – 78) | Tissues |
| LM_CRC | 13 | Sex = male, primary tumor = CRC | N/A | 65 (42 – 77) | Blood |
| Ctrl | 19 | Sex = male, age $\geq 65$ | Cancer/Alzheimer history | 68 (65 – 74) | Blood |

N/A – not applicable

**Table S2. CD4+ T cells and CTLs categorization of the scRNA-seq dataset derived from Zhang et al.,based on patients, tissue type and cell subpopulations (separate Excel file)**

**Table S3.** **The list of top-ranked genes correlated with the expression of *TIGIT* in Tregs located in TME of CRC patients**.

| Gene | Protein | Correlation | Adjusted p-value |
| --- | --- | --- | --- |
| IL32 | Interleukin 32 | 0.31 | $1.82^{-9}$ |
| CD27 | CD27 | 0.30 | $7.92^{-9}$ |
| MAGEH1 | MAGE family member H1 | 0.25 | $7.23^{-6}$ |
| LAYN | Laylin | 0.25 | $1.14^{-5}$ |

**Table S4. The immune profile of an Atlas of cancer cells dataset derived from Nieto et al., distinguished based on patients, cell subpopulations and cancer types (separate Excel file)**

## Notes

### Author Declarations

Independent Bioethics Committee for Research at the Medical University of Gdańsk gave ethical approval number for this work NKBBN/564/2018 with multiple amendments

